# A new strategy on Early diagnosis of cognitive impairment via novel cross-lingual language markers: a non-invasive description and AI analysis for the cookie theft picture

**DOI:** 10.1101/2024.06.30.24309714

**Authors:** Jintao Wang, Junhui Gao, Jinwen Xiao, Jianping Li, Haixia Li, Xinyi Xie, Rundong Tan, Yuyuan Jia, Xinjue Zhang, Chen Zhang, Dake Yang, Gang Xu, Rujin Ren, Gang Wang

## Abstract

**Background:** Cognitive impairment (CI), including Alzheimer’s disease (AD) and mild cognitive impairment (MCI), has been a major research focus for early diagnosis. Both speech assessment and artificial intelligence (AI) have started to be applied in this field, but faces challenges with limited language type assessment and ethical concerns due to the “black box” nature. Here, we explore a new stragety with patient led non-invasive observation for a novel cross-lingual digital language marker with both diagnostic accuracy, scalability and interpretability.

**Methods:** Speech data was recorded from the cookie theft task in 3 cohorts. And automatic speech recognition (ASR), Networkx package, jieba library and other tools were used to extract visual, acoustic and language features. The SHAP model was used to screen features. Logistic regression and support vector machine and other methods were used to build the model, and an independent cohort was used for external verification. Finally, we used AIGC technology to further reproduce the entire task process.

**Results:** In Chinese environment, we built 3 models of NC/aMCI, NC/AD, and NC/CI (aMCI+AD) through Cohort 1 (NC n=57, aMCI n=62, AD n=66), with accuracy rates of 0.83, 0.79, and 0.79 respectively. The accuracy was 0.75 in the external scalability verification of Cohort 3 (NC n=38, CI n=62). Finally, we built a cross-lingual (Chinese and English) model through Cohort 1 and 2, built a NC/aMCI diagnosis model, and the diagnostic accuracy rate was 0.76. Lastly, we successfully recreate the testing process through Text-to-Image’ and Animation Generation.

**Discussion:** The visual features created by our research group and combines acoustic and linguistic features were used to build a model for early diagnosis of cognitive impairment, and a cross-lingual model covering English and Chinese, which performs well in external verification of independent cohorts. Finally, we innovatively used AI-generated videos to show the subject’s task process to the physician to assist in judging the patient’s diagnosis.

**Keyword:** Alzheimer’s disease, Amnestic mild cognitive impairment, speech test, Artificial Intelligence, interpretability

## INTRODUCTION

Alzheimer’s disease (AD) is the most common neurodegenerative disease, characterized by persistent memory decline. It is reported that 55 million^1^ people suffer from AD and other dementia worldwide and 13 million in China^2^. Thus, dementia has been considered one of the global health dilemmas. Detecting individuals who are at the early stage of dementia is essential but challenging, especially in the disease modified therapy (DMT) era of monoclonal antibodies that emphasizes early diagnosis and treatment. In the past few decades, major progress has been made in the development of biofluid or neuroimaging biomarkers for early screening and/or early diagnosis. However, these methods are limited for application by being invasive and/or expensive. On the contrary, previous studies showed that verbal categorical fluency test showed the highest performance in differentiating AD with the heath^3^, and series of studies^4–6^ have demonstrated that language deficits precede memory impairment. Therefore, early diagnosis through language features is feasible^7^. Previous studies from our team and others suggested that cognitive impairments have been effectively diagnosed through acoustic and linguistic features such as percentage of silence duration (PSD)^8–11^. These features come from the picture description task “cookie-theft”, which has become one of the most commonly used tests of language function and was originally part of the Boston Diagnostic Aphasia Examination (BDAE) manual^12^. However, these methods only consider language characteristics and ignore other cognitive abilities involved in describing the task. Screening of cognitive function by past traits may be inadequate, and interpretability is insufficient.

Meanwhile, advances in artificial intelligence (AI) technology have sparked new research interest for easy and even remote detection, diagnosis and treatment of dementia^13^. Natural language process (NLP) models have achieved relatively better prediction accuracy, even higher than 90%^14^. However, clinical medicine relies on the transparency of decision-making, and the logic of black-box models violates medical ethics. Clinicians could not reasonably accept and understand the decision-making process with no explainable AI (black-box models). Therefore, explainable AI has become a hot topic of research in academia, industry, and government. The interpretability of AI in the medical field has received widespread attention due to its high-risk nature. Additionally, few studies have addressed the problem of cross-language screening, and common issues between different languages are difficult to discover. Most studies are limited to the detection of small samples in a single language with difficult reproducibility. In the present, unfortunately, there are no established and widely accepted methods so far^15^.

Therefore, we here construct a novel language-related digital model with both accuracy and interpretability. Especially, the “cookie theft” task in our study was participant-led, without unnecessary prompts of the physician, which can fully reflect their comprehensive cognition rather than only language ability. So, firstly we create a set of new features named visual features, which can describe the entities and the relationship paths between the entities from the speech to reflect the task process. Together with acoustic or linguistic features mentioned in the previous studies, we construct models to distinguish NC (normal control) from aMCI (mild cognitive impairment), NC from CI (cognitive impairment), and NC from AD. Secondly, these new features can solve the cross-language issues which not handled well before^16^ regardless of language type (Mandarin or English). Finally, we use Artificial Intelligence Generated Content (AIGC) to reproduce the task process, allowing physicians to participate in the classification to reduce overall errors in clinical setting.

## METHOD

### 1. Participant

Cohort1: This is a cross-sectional study, with a total of 185 participants recruited from Ruijin Hospital Affiliated to Shanghai Jiao Tong University School of Medicine, Shanghai, in which 57 were NC, 62 were aMCI, and 66 participants were diagnosed with early phase AD. The registration number is ChiCTR2000036718 on the website associated with this study (https://www.chictr.org.cn). All participants were recruited between August 2020 and August 2023 from the memory clinic of Ruijin Hospital. The authors asserted that all procedures contributing to this work comply with the ethical standards of the relevant national and institutional committees on human experimentation and with the Helsinki Declaration of 1975, as revised in 2008. All procedures involving human subjects/patients were approved by the Ethics Committee of the Ruijin Hospital (approval number: 2020-261). All included individuals provided written consent.

Cohort2: In order to build cross-language models and better generalization capabilities, the DementiaBank corpus was kindly used in the present study^17^. This corpus contained recordings of 74 controls and 25 aMCI patients, from July 1983 to April 1988 (last modified in November 2018) involving the participants given a picture description task, which was originally designed for the Boston Diagnostic Aphasia Examination. The task required each participant to describe events depicted in the picture, the same as performed by participants in our center (Cookie Theft picture description task).

Cohort3: To further verify the accuracy of the model, we randomly included 100 external validation cohorts from the Alzheimer’s disease and other dementia clinical cohorts of Ruijin Hospital (Approval number: 2022-097). These include 62 CI patients(MCI due to AD and mild AD) proven by AV45 PET scans and 38 matched cases with normal cognition. The speech task was performed as what has mentioned above.

### 2. Clinical assessment and diagnosis

To exclude other causes of cognitive impairment, we performed cranial MRI or computed tomography (CT) to exclude confounding factors such as stroke or intracranial space-occupying lesions. Serum folic acid, vitamin B12 levels, and thyroid function were tested to exclude endocrine and metabolic disorders. Clinical and demographic data including age, gender, and level of education were also collected. All subjects underwent neuropsychological tests including the Mini-Mental State Examination (MMSE) , Clinical dementia rating scale(CDR) and the Cookie-theft picture description task from the Boston Diagnostic Aphasia Scales.

After clinical assessment, the participants were categorized into three groups: (i) a NC group, who were considered as cognitively healthy after the clinical consultation; (ii) an AD group, whose diagnosis was based on the clinical probable criteria for diagnosis of AD issued by the National Institute on Aging-Alzheimer’s Association workgroups in 2011^18^; and (iii) an aMCI group, in which patients had a memory complaint corroborated by at least one informant, and a diagnosis was conducted using the Petersen criteria^19^. Participants were excluded if they had any other neurological diseases, any systemic disease which can lead to cognitive dysfunction, psychiatric disorders, or severe hearing or vision impairment. 62 of the subjects accepted dual-phase [18F] AV45 PET scans, with a resolution of 3.76 × 3.76 × 4.9 mm^3^ (field of view = 157 mm). Forty-seven planes were obtained with a voxel size of 1.95 × 1.95 × 3.2 mm^3^. A transmission scan was performed for attenuation correction before the PET acquisition. For [18F] AV45 PET, each participant underwent a 10-minutes early acquisition (composed of ten 1-minute dynamical frames) that began immediately after the intravenous injection of ∼ 4 MBq/kg of [18F] AV45, and a 10-minutes late acquisition (beginning 50-minutes after injection).

Subjects with MMSE ≥ 15 was the include according to the same standards as before^8^, and all enrolled patients were aMCI or mild AD patients.

### 3. Recording protocol

Subjects performed a Cookie Theft picture description task, during which they were given a picture and were told to discuss everything they could see happening in the picture in 1 min while being recorded. The mean time duration of the records is 42.87±18.72s. The RSF cohort individuals’ speech was recorded under the following configuration parameters of Cool Edit Pro software: a frequency of 160000 Hz, creating a 16-bit mono recording, and environmental noise was limited to under 45 dB. The Pitt cohort and Cohort 3 records (the mean time duration of the records is 51.1±23.63s seconds and 43.52±18.42s) were converted to the audio configuration parameters identical to the RSF recording using the Cool Edit Pro software.

### 4. Information generation and processing

### 4. Feature Engineering

#### 4.1 Sound Feature Extraction

The automatic speech recognition (ASR) software for cognitive impairment v1.3 (developed by our team, China Software Copyright number 2016SR164680) for speech analysis was used, according to our previous study^8,9^. Each sample was analyzed by ASR software for cognitive impairment using v1.3 to extract the speech/silence parameters. The sum of all silent periods divided by the total speech time is the definition of PSD (ratio of total silent pause duration to total speech duration), expressed as a percentage. The definition of basic parameters set in our software was according to Pakhomov et al. Who had developed the measurements of spontaneous speech from the Cookie Theft picture description task for patients with dementia. Silence is defined as the summed duration of all silent segments of the recording, including general short pauses, general long pauses, and hesitation-associated pauses.

#### 4.2 Speech-to-Text Conversion

Upon obtaining the audio, after confirming its integrity, the software was utilized for conversion, resulting in a transcript. The audio was then replayed to proofread the transcript, with discrepancies from the original audio requiring modification. The revised transcript constituted the final version.

#### 4.3 Part-of-Speech Tagging of Text

For both Chinese and English texts, specific parts of speech (adjectives, adverbs, prepositions, etc.) in the description texts of AD patients were statistically analyzed using the Jieba library. Specifically, by utilizing the Jieba library (the best library for Chinese text processing), text segmentation was performed for Chinese texts, followed by part-of-speech tagging to obtain the part of speech for each word. Subsequently, the total word count and the count of specific parts of speech were calculated to determine the proportion of each part of speech in the text. Analyzing the distribution of specific parts of speech in the given text aids in understanding the linguistic characteristics of the text.

#### 4.4 Visual Feature Extraction Construction of Spatial Semantic Graphs

Initially, the two-dimensional centroid coordinates (xi, yi) of entities were calculated, and spatial semantic graphs were constructed for each participant. For each participant, the text was scanned from start to finish to extract entities, obtaining an entity list. A directed graph was then constructed based on the entity list and their coordinates.

##### Feature Extraction of Spatial Semantic Graphs

Networkx (v2.8.4 https://github.com/networkx/networkx) were utilized to extract features from graphs. Representative strength features include: the number of edges, path length, number of left-right switches, and diameter of the gaze area. Features representing efficiency include entity density. Features representing attention include: the number of loops, proportion of left-right descriptions in the graph, and graph density.

### 5. Model Construction

#### 5.1 Feature selection as the digital markers

SHAP (SHapley Additive exPlanations v0.44.0)^20^ was used to explain the predictions of machine learning (ML) models based on the Shapley value concept from cooperative game theory. Shapley values measure each participant’s contribution to the outcome of the game, and in ML, they are used to quantify each feature’s impact on the model’s output.

At the same time, based on the assumption that language impairment will worsen with cognitive impairment, only features with consistent trends among NC, aMCI, and AD groups will be included as digital markers and ranked according to Shapley values. Specifically, Random Forest Classifier was chosen as the ML model and fitted on the training set. Predictions were made on the test set, and the model’s accuracy (accuracy_score) was computed. Then, an explainer was created using the SHAP library to calculate and visualize Shapley values, explaining the model’s contribution to the predictions. Finally, force_plot was used to display Shapley values for the certain sample.

#### 5.2 Construction of Single-Language Models

We performed the construction of three models for NC (n=57) and aMCI (n=62), NC (n=57) and AD (n=66), and NC (n=57) and CI (AD+aMCI) (n=128), respectively. The data were partitioned into training and testing sets using the train_test_split function in Sklearn package (v1.0.2)^21^, with 80% allocated for training and 20% for testing to evaluate model performance. Feature filtering and selection included accurate evaluation of features’ consistency with the disease, balance among three factors (expression intensity, expression efficiency, attention), etc. High-credibility models recognized in clinical research were selected: logistic regression (LR), support vector machine (SVM), random forest (RF), k-nearest neighbors (KNN), etc.

#### 5.3 Construction of Cross-Language Models

This study presents an analysis of classification performance on a dataset comprising 218 samples of mixed-language texts from both normal individuals (NC) and those with amnesiac mild cognitive impairment (aMCI). The dataset encompasses 8 lexical diversity features, 9 visual features, and 6 pause-related features. The data were partitioned into training and testing sets using the train_test_split function, with 80% allocated for training and 20% for testing to evaluate model performance. A logistic regression model was initialized and fitted to the training set. The fitted model was then used to predict on the testing set, yielding probability values. These probabilities were utilized to compute parameters of the Receiver Operating Characteristic (ROC) curve, including True Positive Rate (TPR) and False Positive Rate (FPR) for model evaluation. In the graphical representation, the orange curve represents the ROC curve, with the Area Under the Curve (AUC) serving as one of the metrics for evaluating model performance. Higher AUC values signify superior model performance.

### 6. Process Reproduction

#### 6.1 Image Generation

Our method is based on the open-source project stable-diffusion-webui v4.7^22^, which implements Stable Diffusion, a deep learning model for image generation. We introduced the Control Net plugin (v11p-sd15)^23^ into this method, which extends the functionality of Stable Diffusion to better control the layout and content of images. Specifically, we used Text-Guided and Image-Guided functions to redraw the generated images. The Text-Guided function allows us to use textual descriptions to guide the image generation process, such as specifying the theme or content direction of the image. The Image-Guided function allows us to use reference images to guide image generation, ensuring consistency in content and style between the generated image and the reference image. In summary, our method combines Stable Diffusion algorithm with the ControlNet plugin, as well as Text-Guided and Image-Guided functions, to achieve fine control and redraw of the image generation process, thereby generating high-quality images with specific content and layout. Specific to our language description, after inputting the content of the text conversion, we can obtain high-quality restored pictures.

#### 6.2 Animation Generation

Same as the Image Generation process, iterating through the text describing the images by participants, starting with a blank image, a new image was generated if the number of entities increased. Finally, all images were concatenated in the order they were generated to create the animation. Here, the duration of each image corresponds to the time in the original audio

### 7 Statistical analysis

The demographic information analysis was performed using SPSS (Version 26.0). T test and one-way ANOVA were used for the group differences, Turkey test was used for post-hoc analusis. Chi-square test and Fisher test were used to detect the frequency differences between groups.The Pearson correlation was used for the association between features and MMSE subscore. *P* < 0.05 were considered significant

## RESULTS

### 1. Demographic information

Cohort1:

There were 57 NC, 62 aMCI, and 66 AD patients in the cohort1. Gender (female in NC: 59.65%, aMCI: 58.06%, and AD: 48.48 %) and educational level (NC: 14.98 ± 3.15 years, aMCI: 14.76 ± 3.18 years, and AD: 14.68 ± 3.51 years) showed no significant difference among the NC, aMCI, and AD groups in the cohort, and mean age was 69.60 ± 7.71, 72.82 ± 7.43, and 73.21 ± 8.63 years for the NC, aMCI, and AD groups within this cohort (P= 0.0269), respectively. However, there were significant differences between groups’ mean MMSE scores (NC: 29.11 ± 0.99, aMCI: 25.64 ± 4.31, and AD: 18.85 ± 3.38), and subitems relating to each cognitive domain (Table 1, all P < 0.0001), and the post-hoc comparison results are shown in Table 1.

**Table 1.**
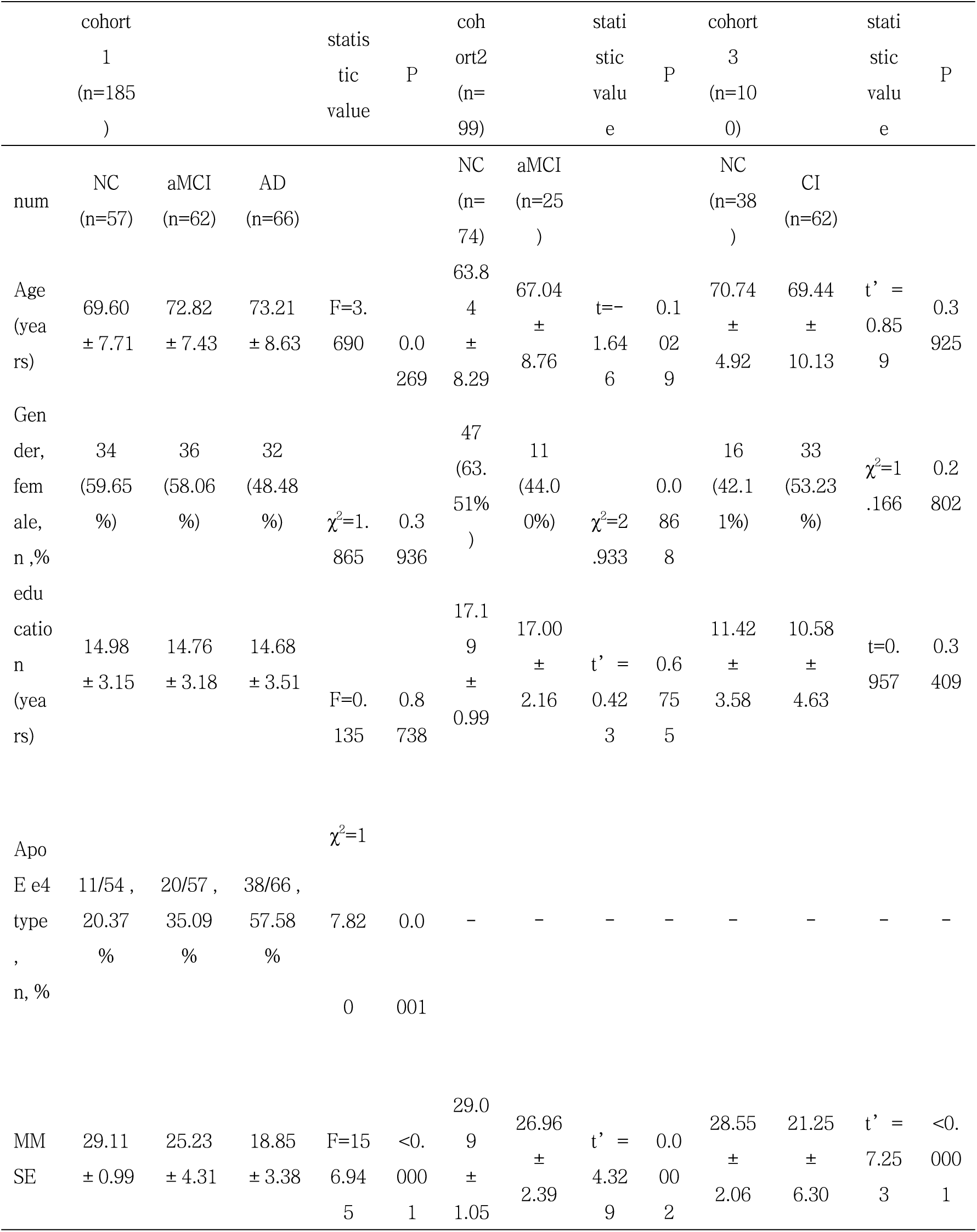
The demographic feature of subjects.

Cohort2:

There were 74 NC, 25 aMCI patients in the Pitt cohorts. Gender (female makeup in NC: 63.51%, and aMCI : 44.00%) and educational level (NC: 17.19 ± 0.99 years, and aMCI : 17.00 ± 2.16years)s, and mean age (63.84 ± 8.29, and 67.04 ± 8.76 years for the NC, and aMCI groups) showed no significant difference among the NC, and aMCI group (P>0.05). There were significant differences between groups’ mean MMSE scores (NC:29.09 ± 1.05, and aMCI :26.96 ± 2.39) (P=0.0002).

Cohort3:

There were 38 NC, 68 CI patients in the Alzheimer’s disease and other dementia clinical cohorts. Gender (female makeup in NC: 42.11%, and CI : 53.23%) and educational level (NC: 11.42 ± 3.58 years, and CI : 10.58 ± 4.63 years) , and mean age (70.74 ± 4.92, and 69.44 ± 10.13 years for the NC, and CI) showed no significant difference among the NC, and CI groups (P>0.05). There were significant differences between groups’ mean MMSE scores (NC: 28.55 ± 2.06, and CI : 21.25 ± 6.30) (P <0.0001).

### 2. Feature selection as the digital markers

We ultimately selected 23 features as the digital markers for cognitive impairment related to the subjects’ voice, language, and vision. Their IDs and descriptions are shown in the table below.

**Table 2.**
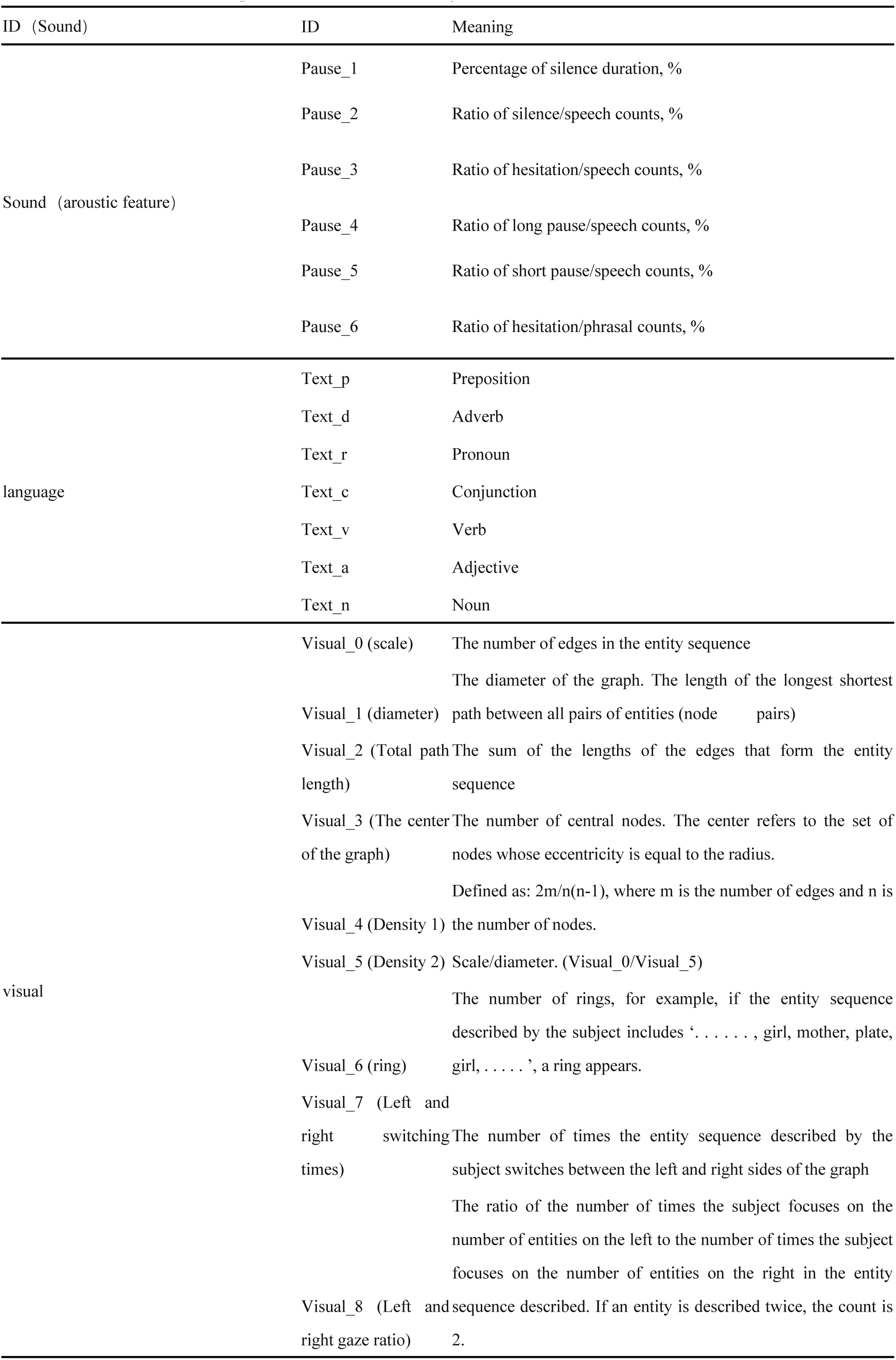
The IDs and Descriptions of the Three Categories of Features.

We randomly selected the SHAP values for three samples Figure 1. According to the plots, the contribution value of each feature influences the model’s prediction for a specific sample. The plot shows how the model’s prediction for a sample is composed of the impacts of individual features. In these samples, pause features (Pause_1-3) and the proportions of specific parts of speech (mainly nouns and prepositions) play a significant role in the model’s predictions and how they influence the model’s decisions. By calculating the average SHAP values across the three samples, we determine the importance of each feature. Pause_1, Text_n, and Text_p were the most important features in the model. Together with other features, they were included as the digital markers.

**Figure 1.**
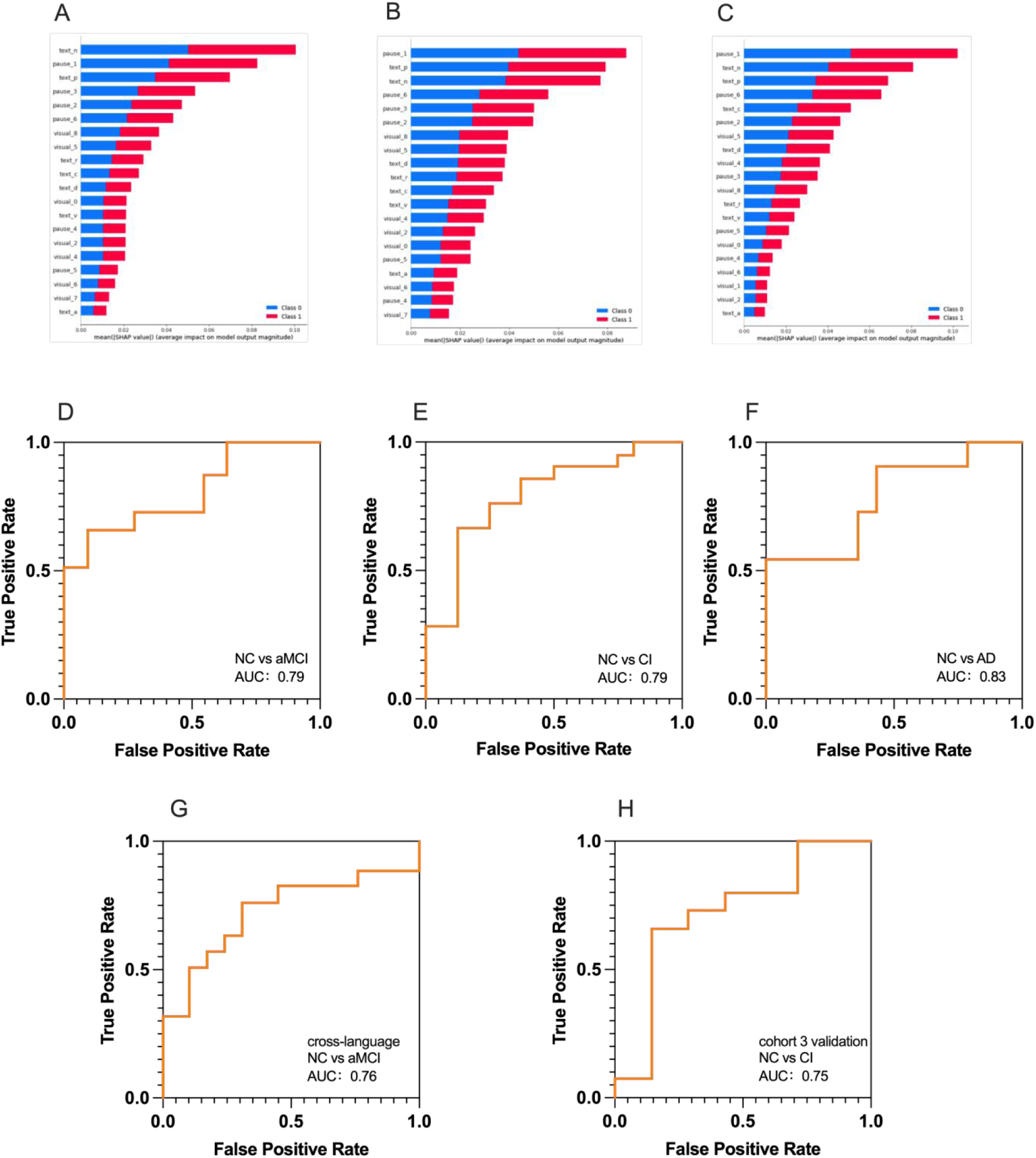
(A-C) The contribution value of different features to prediction of randomly selected three samples, (D-F) Diagnostic model for NC/aMCI, NC/AD, NC/CI. And the area under the curve. (G) In a cross-language environment, the diagnostic ROC curve of diagnostic model for NC/aMCI,and the area under the curve.(G) Validation of the NC/CI diagnostic model in an external cohort 3 and the area under the curve.

**Table 3.** Feature Importance Ranking.

|  | Sample1 | Sample2 | Sample3 | Average |
| --- | --- | --- | --- | --- |
| Pause_1 (PSD) | 2 | 1 | 1 | 1.33 |
| Text_n (Noun) | 1 | 3 | 2 | 2.00 |
| Text_p (Preposition) | 3 | 2 | 3 | 2.67 |
| Pause_6 (hesitation counts) | 6 | 4 | 4 | 4.67 |
| Pause_2 (RSD) | 5 | 6 | 6 | 5.67 |
| Pause_3 (hesitation ) | 4 | 5 | 10 | 6.33 |
| Visual_5 (Density 2) | 8 | 8 | 7 | 7.67 |
| Visual_8 ( Left and right gaze ratio) | 7 | 7 | 11 | 8.33 |
| Text_d (Adverb) | 9 | 9 | 8 | 8.67 |

### 3. Employing Machine Learning Methods Accepted by Clinicians

A ML model was constructed to perform a binary classification between NC/aMCI, NC/AD, NC/CI (aMCI+AD), The ROC curves comparing PSD based classifcation sensitivity and specifcity among NC, aMCI, and AD patients are shown in Fig.1A–D. The AUCs of the curves are 0.83, 0.79, and 0.79 in NC/aMCI, NC/AD, and NC/CI (aMCI+AD). Further, the sensitivity and specificity of NC/aMCI, NC/AD, and NC/CI (aMCI+AD) is 0.71/0.71, 0.84/0.70, and 0.78/0.79 respectively. The results are shown in the following figure. And the weight of each features were listed in the Supplementary Table 2.

### 4. Constructing Cross-Lingual Models for Further Interpretability Support

The visual features are related to the described order during the examination, and are independent of the language, wording, and sentence structure. We use visual features to solve cross-language diagnostic problems losslessly. In order to clarify the suitable features of the model, all the features need to simultaneously satisfied the ’consistency’ of change in both Chinese and English, which the mean value of this indicator satisfies a monotonic change across the three groups: NC > aMCI > AD, or NC < aMCI < AD. Ultimately, the cross-lingual model achieved an accuracy of 0.76 and a sensitivity of 0.75.

**Figure 2.**
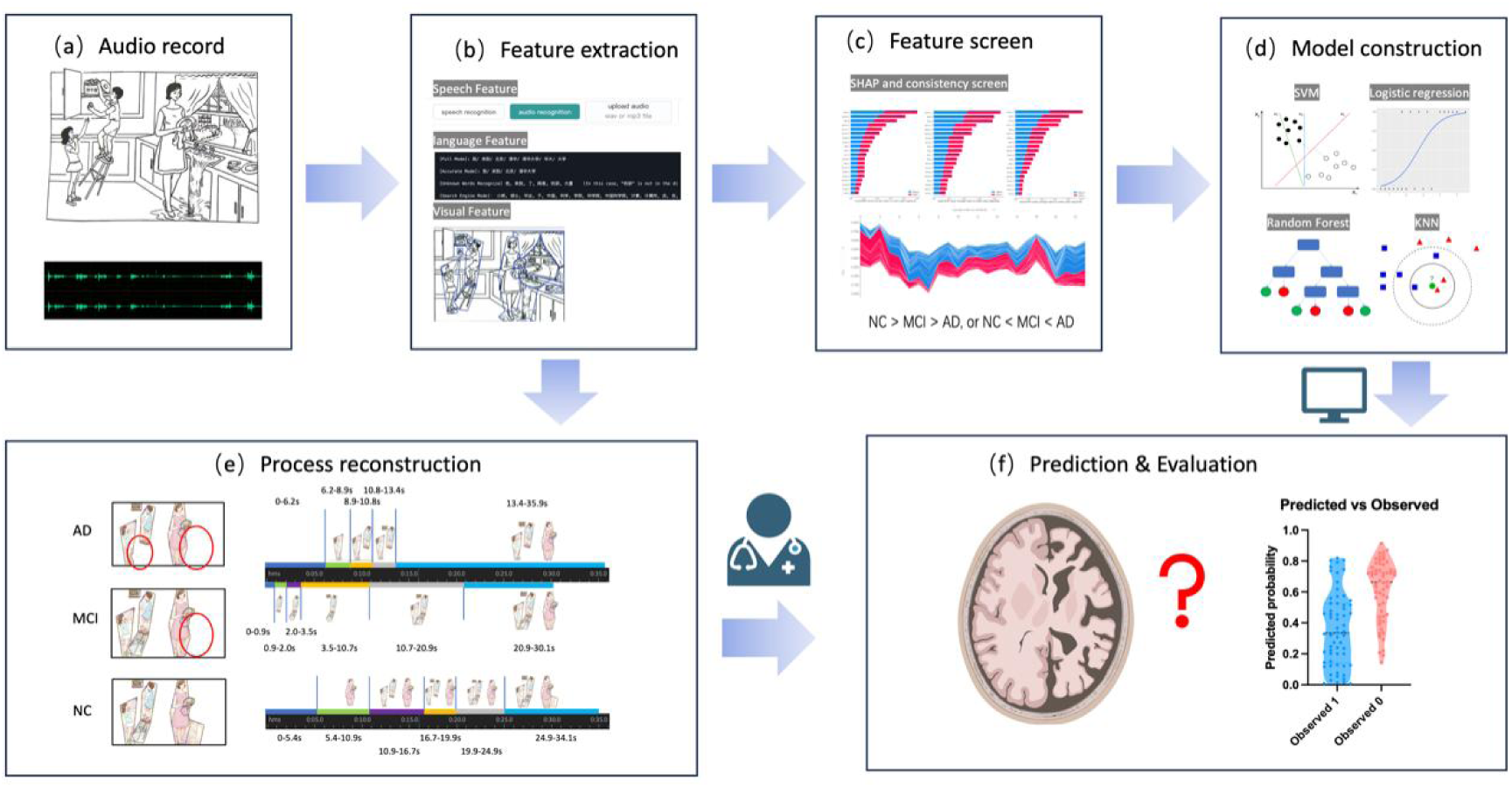
Language Cognition Screening and Reproduction Flowchart

### 5. External validation of machine learning models

To evaluate the effectiveness of our model in distinguishing NC from CI, we used an independent external cohort, and all CI patients in this cohort underwent AV45-PET examination and were confirmed to be Aβ positive. After external validation, the model’s prediction accuracy reached 75%, with sensitivity and specificity of 68.24% and 73.33%.

### 6. association of visual feature and cognitive domain

From the single language model, the visual_0 and visual_4 has the biggest weight for the model, we found they are significantly associated with the MMSE attention and delayed memory subscore(visual_0 with memory: R^2^=0.03891, P=0.0492, visual_4 with memory: R^2^=0.1451, P<0.0001, visual_4 with attention: R^2^=0.09499, P=0.0018). Supplementary Table 1 and **Figure 3**.

**Figure 3.**
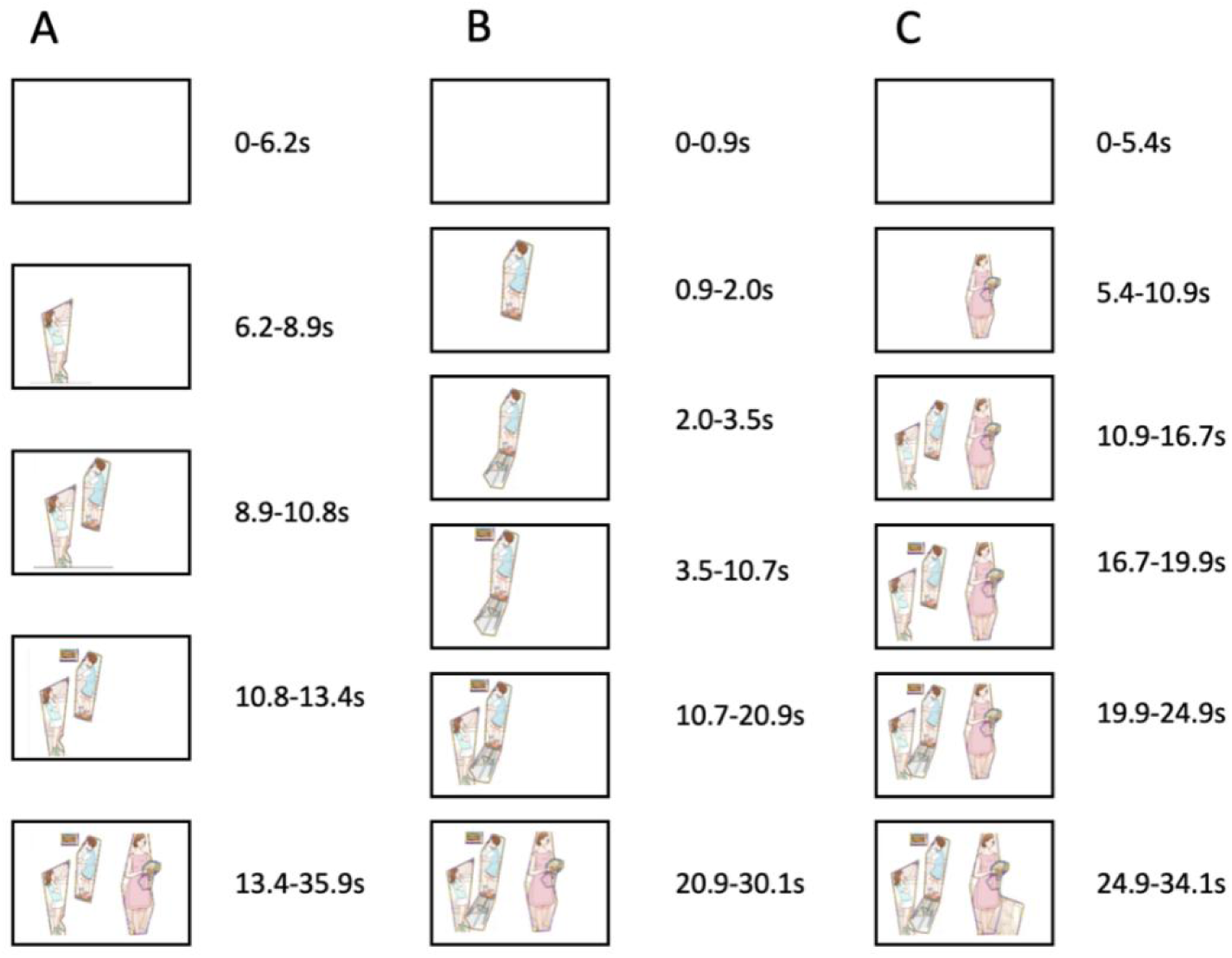
Generated animation screenshot of different timelines. (A-C) the animation of AD, aMCI, NC respectively

### 7. Recreating the Testing Process Through Text-to-Image’ and Animation Generation

The first method for process recreation is “Text-to-Image” to gain the richer content and more cartoon-like features image compared to the other AIGC for example Image-to-Image method. However, the “Text-to-Image” method cannot show the patient’s task description at each time point, and may miss some information. Most importantly, this method is not reproducible and cannot be compared across multiple samples. Therefore, we further used animation generation to completely display the process. The original picture was segmented, and was divided into different entities, which will present according to the task process. The generated animation can inform the doctor which entities were described, which were not described, and the duration of each entity’s description.

## DISCUSSION

In the present study, we successfully constructed a strategy with both accuracy and interpretability, and innovatively created new digital language markers(Figure4). Here, the cookie-theft task was participant-led, and the physician’s role was minimized, providing only necessary prompts to fully reflect their cognitive integration ability. Therefore, we draw on the entity path diagram (EPD) of graph theory to create visual features that could reflect information including language but not limited to language. In addition, with visual features we constructed a cross-language cognitive screening model successfully. Finally, this study innovatively used AIGC to more intuitively cooperate with clinicians in clinical applications through generative images or videos. Regarding to good cost performance and easily handling compared to traditional body fluids and neuroimaging biomarkers, we believe new digital markers have better accessibility advantages. It may be particularly suitable for early screening in multilevel referral systems for cognitive impairment, and particularly in less well-resourced or remote regions.

**Figure 4.**
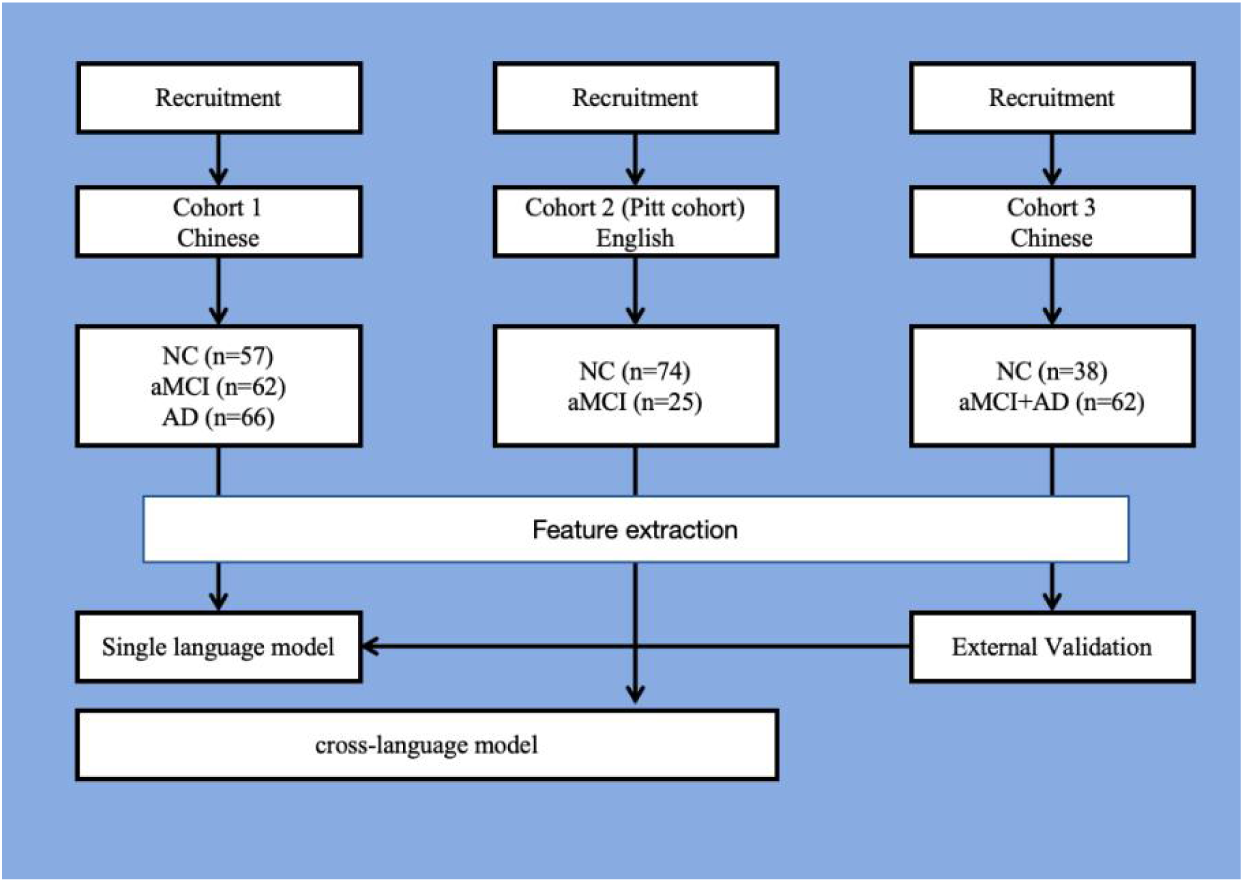
Study design of the clinical trial

### 1 Models via SHAP method with better accuracy and interpretability reduce black box effect confusion

This study adheres to the principle of interpretability in feature selection, and selects interpretable features related to speech, text, and vision through SHAP^20^ to build a classification model. This approach follows the rigor of medical research and ensures that the features used can be used as digital or biomarkers. For our cognitive screening in the Chinese language environment, we constructed classification diagnosis models with accuracy of 0.83, 0.79, and 0.79 in NC/aMCI, NC/AD, and NC/aMCI+AD through ML models such as SVM. We obtained a good prediction accuracy, compared with other previous studies^11,14,25–27^. But unlike those technologies based on large language models, including word2vec, Bert, and GPT, the use of SHAP and SVM makes our model more interpretable rather than an unknown black box process. To further verify the reliability of our model, we introduced an independent cohort 3 for external validation in which all patients had abnormal β amyloid deposits through AV45 PET scans. In cohort 3, our model also achieved a prediction accuracy of 75%, indicating that our model has high external scalability and high value in the diagnosis of cognitive impairment. We further analyzed the weight of each feature in the model of NC/aMCI+AD. Among them, the acoustic features headed by ratio of hesitation/phrasal counts, and PSD discovered by our previous research. The Scale among the visual features also have a greater contribution in this model. We believe the pauses in the speech can well reflect the patient’s cognitive ability^8,9^. A larger PSD indicates worse cognitive function. Scale can refer to the hierarchy or complexity of the graph. In our model, the smaller the complexity, the more likely the patient to be cognitively impaired. People with cognitive impairment describe the task more simply. In addition, we hold that visual features can reflect more cognitive domains than speech ability. We found that visual features with high weights in the model such as visual_0 and visual_4 are closely related to MMSE subscores, especially to cognitive domains such as memory and attention, which further proves the reliability of visual features in cognitive screening.

### 2 Visual features solve cross-language problems well

At present, the biggest barriers in the methods for the speech detection of cognitive impairment is the fact that: most models are language dependent. For example, Yan and colleagues relied on semantic features and existence of transcription tools from any language to English and/or powerful NLP models^28^. Most of these methods convert samples of different languages into the same language, or only use acoustic features for model construction^14^. Translation may lose real information, and the black box properties of the NLP model are also limited. The visual features are less dependent on language type and more related to task completion. Therefore, with the same speech collection conditions, we merge the English speech samples from the Pitt database^17^ and the Chinese speech data in our cohort to truly classify the cognitive levels of different language environments in the same model. We constructed a cross-language model with an higher accuracy of 0.76 compared with 0.70 in Fraser and colleagues’ model^16^ in classifying a mixed sample of Swedish and English. Compared with in single language environment, the model accuracy was similar which means the important role of visual features in these model.

### 3 Image and Animation Generation

In picture description tasks, many studies have implemented relatively automated screening processes, but often a high degree of automation may lead to potential errors that are difficult to detect by clinicians. Therefore, we innovatively made use of AIGC technology combined with visual features to generate images/videos that can reproduce the completion process of language tasks. The images/videos will facilitate more intuitive inspection by clinicians. Information such as the completeness of the content or the intensity of the description, the order of the description, and the spacing will be fully displayed in the picture. In our three patient examples of NC, aMCI, and AD(Figure 3), different patients presented different completion results and different processes, which very intuitively shows the completion status of patients with different cognitive or language abilities.

### 4 Limitation

First, although, we have considered confounding factors such as education level, age, and gender in the model construction, however, language as a high-level brain function is still affected by factors such as religion, emotion, personality traits, and language habits^6,15^. In the future, these factors should be taken into consideration. Secondly, How to avoid the learning effect after multiple tests and reduce the deviation is the direction we are working for. Like other cross-sectional studies^10,29,30^, the conclusions of this study do not have the highest evidence value, and we need more efforts to construct longitudinal cohorts to obtain more reliable conclusions. Thirdly, the etiology of MCI is a heterogeneous^31^. Thus, it is envisioned as future work the implementation of multilingual or language independent systems, supported by extensive and diverse databases (that still must be gathered, with genders, ages, disease severity), as well as the automation of the features selection and extraction. Better decision models, task oriented, are also required.

Generally, the present study successfully creates new digital tool from a new perspective, and uses digital markers and AI to further improve the ability to diagnose early cognitive impairment across languages. Meanwhile, as a vision-related parameter, it can also reflect advanced cognitive functions such as attention and observation. Therefore, our results suggest that visual features can be used not only to screen for cognitive disorders such as AD, but also for diseases related to cognitive changes such as attention deficit and hyperactivity disorder, depression. With the rapid rise of AI, its application has immeasurable prospects. Making full use of new digital markers to diagnose patients with early cognitive impairment will be simpler and more efficient than traditional methods in future.

## Data Availability

All data produced in the present study are available upon reasonable request to the authors

## ACKNOWLEDGEMENT

Sincere thanks to each subject participating in this clinical trial.

## FOOTNOTES

Jintao Wang, Junhui Gao contributed equally.

## CONTRIBUTORS

Conceptualisation—Gang Wang, Jintao Wang, Junhui Gao. Methodology—Junhui Gao, Jintao Wang, Rundong Tan, Yuyuan Jia, Xinjue Zhang, Chen Zhang, Dake Yang, Gang Xu, Rujin Ren, and Gang Wang. Investigation— Jintao Wang, Jinwen Xiao, Jianping Li, Haixia Li, Xinyi Xie,and Gang Wang.

Visualisation—Jintao Wang, Junhui Gao, Rundong Tan, Yuyuan Jia, Xinjue Zhang, Chen Zhang, Dake Yang, and Gang Wang. Funding acquisition—Gang Wang. Project administration—Gang Wang. Supervision—Gang Wang. Writing (original draft)— Gang Wang, Jintao Wang and Junhui Gao. Writing (review and editing)—Gang Wang,Gang Xu and Rujin Ren. Guarantor—Gang Wang.

## SUPPLEMENTAL MATERIAL

